# Comparing the age and sex trajectories of SARS-CoV-2 morbidity with other respiratory pathogens points to potential immune mechanisms

**DOI:** 10.1101/2021.01.07.21249381

**Authors:** C. Jessica E. Metcalf, Juliette Paireau, Megan O’Driscoll, Mathilde Pivette, Bruno Hubert, Isabelle Pontais, Derek Cummings, Simon Cauchemez, Henrik Salje

## Abstract

Comparing age and sex differences in SARS-CoV-2 hospitalization and mortality with influenza and other health outcomes opens the way to generating hypotheses as to the underlying mechanisms, building on the extraordinary advances in immunology and physiology that have occurred over the last year. Notable departures in health outcomes starting around puberty suggest that burdens associated with influenza and other causes are reduced relative to the two emergent coronaviruses over much of adult life. Two possible hypotheses could explain this: protective adaptive immunity for influenza and other infections, or greater sensitivity to immunosenescence in the coronaviruses. Comparison of sex differences suggest an important role for adaptive immunity; but immunosenescence might also be relevant, if males experience faster immunosenescence. Involvement of the renin-angiotensin-system in SARS-CoV-2 infection might drive high sensitivity to disruptions of homeostasis. Overall, these results highlight the long tail of vulnerability in the age profile relevant to the emergent coronaviruses, which more transmissible variants have the potential to uncover at the younger end of the scale, and aging populations will expose at the other end of the scale.

## Introduction

From early in the pandemic, the discrepancy in SARS-CoV-2 outcomes across age was striking (1); evidence for sex differences followed soon afterwards (2). Age profiles associated with infectious disease contain the signature of both transmission dynamics and vulnerabilities to infection (3). For our analysis of emerging pathogens, and influenza where waning of immunity occurs, we consider the variability in risk of infection across age to be secondary to variability associated with health vulnerabilities. We use age and sex profiles of health outcomes in the context of known features of immune function to probe the underlying mechanisms of the morbidity and mortality impact of SARS-CoV-2.

## Results and Discussion

We analysed age and sex data for hospitalization, severe disease (requiring intensive care or resulting in death) and death for SARS-CoV-2 (France and globally), MERS (Saudi Arabia), SARS (China), influenza (France and US), as well as all-cause hospitalization (France) and mortality (28 European countries) (Figure S1). For each pathogen, we compare the incidence within each age-group with the incidence observed in 60 year olds (Figure A-C, Figure S2). This approach allows us to adjust for differences in mortality that affect all age groups and the age structure of the underlying population. We also compare the incidence between males and females (Figure D-F, Figure S2).

The relative increase in the probabilities of hospitalization, severe disease and death are strikingly consistent for older individuals across pathogens as well as with all-cause hospitalization and mortality. For COVID-19 cases over 40y in age, each additional year of life was associated with a 10.7% (95%CI: 10.3-11.0) relative increase in probability of death. This was very similar to SARS (10.2%, 5.2-15.5), MERS (7.1%, 6.2-8.0), influenza (8.5%, 8.1-8.8) and all cause mortality (10.2%, 10.1-104%), and, indeed, the latter has been previously noted (4, 5). Influenza age distributions were also consistent across countries and across different reporting systems within the same country (Figures S3-S4). However, there are notable departures between pathogens among younger age groups.

Qualitatively, all age-patterns echo the classic U-shape of human mortality (6): initially high levels of infant hospitalization, severe disease and death first fall, and then increase (Figure 1A-C). However, the age at which the increase in hospitalization, severe disease or mortality occurs differs considerably. In particular, for both influenza and all cause hospitalisations, rates remain relatively low between ages 10 and 50, while a stark increase is observed for SARS-CoV-2 right after the minima of 9; and for MERS from 6 years (Figure 1A). Severe disease (Figure 1B) and deaths (Figure 1C) also exhibit a steeper increase from a younger age for the coronaviruses, although the long level period is less apparent.

**Figure.**
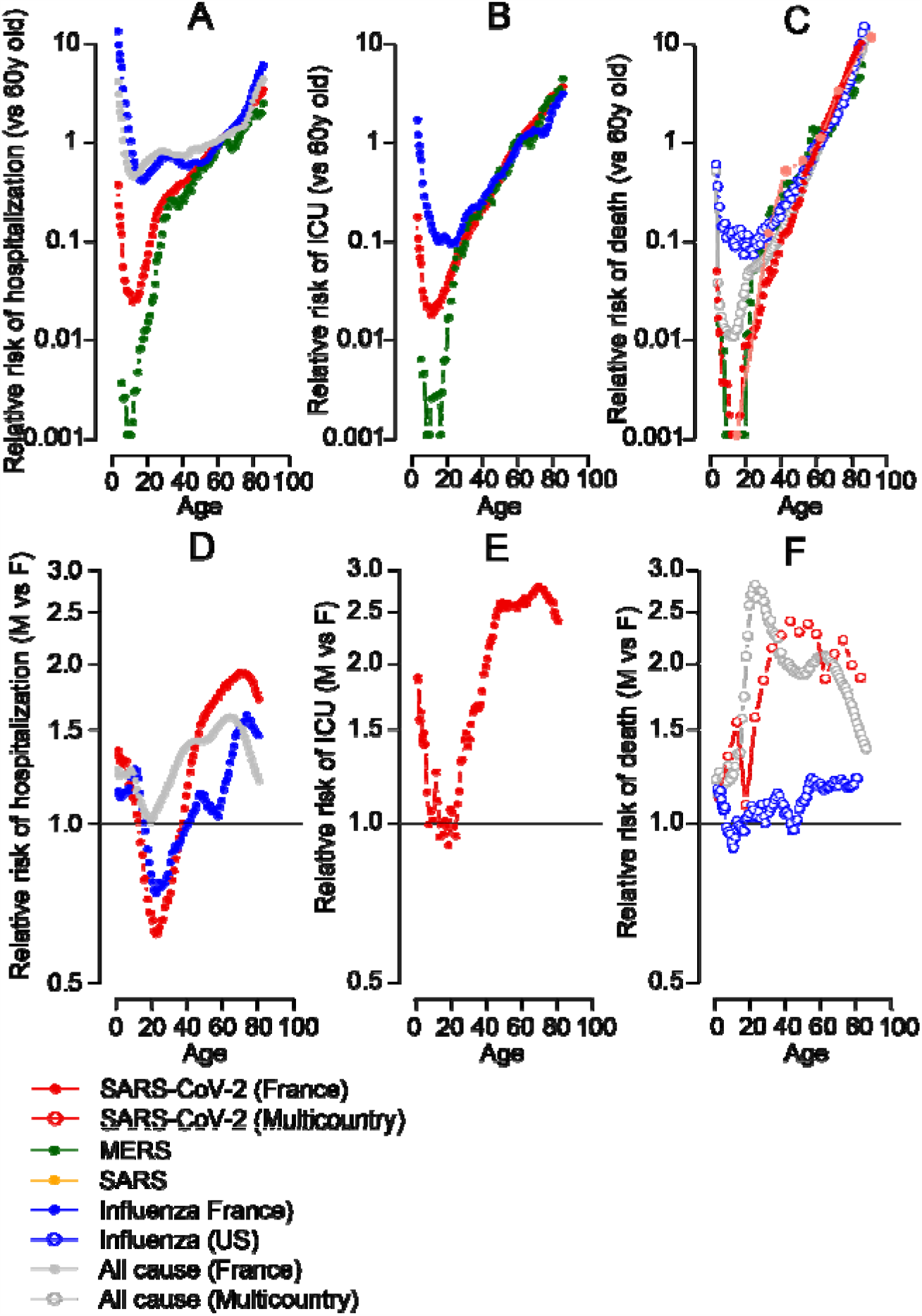
Relative risk of hospitalization, ICU or death for different coronaviruses, influenza as well as underlying levels for all causes. **(A)** Relative incidence of COVID-19 (red), MERS (green), influenza (blue) hospitalization for each age as compared to 60 year olds. The relative incidence of all emergency visits is shown as a comparison (grey). **(B)** Relative incidence of COVID-19 (red), MERS (green), influenza (blue) hospitalization requiring ICU and/or resulting in death for each age as compared to 60 year olds. **(C)** Relative incidence of COVID-19 (red), MERS (green), influenza (blue), SARS (red) deaths for each age as compared to 60 year olds. **(D)** Relative incidence of COVID-19 (red) and influenza (blue) hospitalization for each age comparing males with females. The relative incidence of all emergency visits is shown as a comparison (grey). **(E)** Relative incidence of COVID-19 hospitalization requiring ICU and/or resulting in death for each age comparing males with females. **(F)** Relative incidence of COVID-19 deaths likewise.

These patterns will be shaped by immune functioning over age (7). Early in life, development of immune capacity and memory increase protection from infectious diseases. These gains are then eroded by immunosenescence (8): dysregulation of innate immunity and depletion of adaptive effectors including naive B and T-cells diminishes defense against infectious pathogens or cancer, and increases inflammation (9). The latter effect, sometimes termed ‘inflammaging’ (10), is associated with diseases of old age (cardiovascular disease, strokes, diabetes, etc). The remarkable consistency of late life increases in hospitalization, ICU and death across pathogens (Figure 1A-C) suggests a common mechanism, and immunosenescence is the most obvious candidate. But what do discrepancies earlier in life suggest?

Theory predicts that immunosenescence should begin at the onset of puberty (11), and indeed, immunopathology-associated cytokines (like IL-6) gradually increase with age in healthy adults (12, 13). Falling levels of protective cytokines (like IL-17A and Ifn-gamma) across pediatric and adult SARS-CoV-2 patients (14) echo this result. Thus, one interpretation of the early start to the increase in hospitalization, severe disease and death in SARS-CoV-2 and MERS is that coronaviruses are more sensitive than influenza to this background immunosenescence at earlier life stages.

Two possible mechanisms can be invoked to explain this. First, children are likely to experience several influenza infections early in life (15), and may accumulate sufficient immunity to buffer them from either infection or its consequences for much of adulthood. This is not the case for the two emergent coronaviruses, SARS-CoV-2 or MERS. Second, coronavirus dependence on the ACE2 receptor might make immunopathology a particular risk even for relatively minor viral loads (16), as the ACE2 receptor acts to reduce inflammation. The resulting sensitivity to disruption of homeostasis could interact with the age trajectory of immune decline, so that some threshold for severe outcomes was crossed earlier in coronaviruses than in influenza. The first explanation is thus rooted in development of adaptive immunity, while the second is associated with more general disruption of immune functioning starting around puberty which the emergent coronaviruses might be more likely to trigger.

Sex differences in immunity (17) provide another lense into evaluating possible mechanisms underpinning the burden of SARS-CoV-2. In both influenza and SARS-CoV-2, the burden of hospitalization shifts from being male-biased at the youngest ages, to female between the ages of 18 and 24 and then back to male again at late ages (Figure 1D), with a slight late life reduction in relative male burden at very late ages presumably attributable to frailty effects (18). This broad trajectory is also seen for severe cases of SARS-CoV-2 (Figure 1E). Mortality data lack these non-linearities, and the male burden grows consistently relative to the female burden in both influenza and SARS-CoV-2 across age, but this increase is much more rapid in SARS-CoV-2 (Figure 1F).

Typically, a male bias in risk occurs when weak immune responses underlie significant host damage by a pathogen, while a female bias occurs when strong immune responses promote host damage (19). Respiratory infections typically trigger strong Type 1 immune responses, resulting in inflammation-associated damage to the lung. As estrogens are associated with efficient immune recruitment and response (20–22), while androgens are largely characterized as immunosuppressive (17), greater risk of hospitalization in females after puberty and before menopause is not unexpected for respiratory pathogens like SARS-CoV-2 and influenza (Figure 1D), with further potential amplification of vulnerability during pregnancy (although this is generally only up to 5% of the population hospitalized)(23). Interestingly, however, this greater risk of hospitalization does not translate into much greater risk of mortality in females for either pathogen (Figure 1F), echoing the ‘male-female health survival paradox’, (where females in worse health survive better than comparable males (24)) although this effect is typically reported at much later ages.

Advances in SARS-CoV-2 immunology broadly aligns with expectations for sex differences. Greater male pathology seems associated with a failure to control the virus soon after infection (16, 25–27), while females may both detect (28, 29) and respond (30, 31) earlier. This female advantage may compound over age as a result of the age trajectories of particular immune effectors, e.g., Mucosal Associated Invariant T-cells which are an important line of defense in the lungs recruit more efficiently to airways (32), and deplete less rapidly with age in female blood. Conversely, female severe outcomes seem driven by over-responding immune systems, i.e., higher levels of innate immune cytokines were associated with worse disease progression in females, but not males (27). Assuming that the broad general patterns hold, for influenza, adaptive immunity could benefit females by diminishing excessive immune responses, or males by enabling more efficient pathogen control. If influenza and SARS-CoV-2 are otherwise comparable, hospitalization data suggests that immunity benefits females, as the hospitalization data indicates a reduced bias towards female burden in influenza relative to SARS-CoV-2 for ages <30 years (blue line above the red line, Figure 1D); the bias then moves into males in SARS-CoV-2 at a younger age than for influenza, and accelerates more rapidly (Figure 1D), also observed for mortality (Figure 1F). However, although these patterns suggest that immunity could explain the age and sex differences in SARS-CoV-2 and influenza, immuno-senescence cannot be dismissed faster male immunosenescence (33, 34) could also drive these results.

Despite the fact that younger individuals have been less vulnerable to the pandemic virus to date, probing the age and sex trajectories of SARS-CoV-2 suggests that a lack of protective immunity, potentially amplified by the intersection of viral receptor use with host homeostasis mechanisms can expose an early start to immunosenescence. Viral evolution might expose this, which would increase the morbidity and mortality burden associated with the pandemic virus; which vaccination might provide an opportunity to buffer.. More generally, as the global population ages, the intersection of immuno-senescence with pathogens for which we have no adaptive immunity may open important vulnerabilities to emergent pathogens.

## Data Availability

Data is available on request to the authors.

## Materials and Methods

### Data sources

#### 1. COVID-19 in France

We obtained hospitalisation data from the French SI-VIC system from the period March-September 2020. The SI-VIC system is integrated into all hospitals in France and has been used since March 2020 to monitor COVID-19 hospitalised patients. Data are sent daily to Santé publique France, the French national public health agency. We obtained the number of hospitalisations, the number of patients requiring intensive care treatment and the number of patients that died for single year age classes and by sex.

#### 2. Influenza emergency visits in France

We obtained the age and sex distribution of emergency visits from the French OSCOUR® network between 2004 and 2019, where influenza was identified as the main or associated clinical diagnosis(35). The OSCOUR® network was set up in 2004 by Santé publique France and now covers about 95% of all French hospitals. We removed data from 2009 from this dataset as 2009 was a pandemic year with different age-distribution of cases.

#### 4. Influenza ICU and death data in France

We obtained the number of cases requiring ICU and the number of deaths by age for the period 2012-2017, from the French national hospital discharge database (PMSI), which contains all records of hospitalisations in all hospitals in France (36). We included hospital stays where influenza was identified as the main or associated clinical diagnosis.

#### 5. Influenza mortality data USA

We extracted the number of deaths by single year age class and by sex where influenza was the main cause of death for the whole of the USA in 2019 from the US National Vital Statistics System (https://www.cdc.gov/nchs/nvss/deaths.htm).

#### 6. MERS data

We used the MERS database maintained by Andrew Rambaut at the University of Edinburgh that was collated during the MERS outbreak in 2013 (https://github.com/rambaut/MERS-Cases/). This linelist database province individual age, sex and whether the individual required intensive care treatment and whether they died.

#### 7. SARS mortality data

We obtained the number of SARS deaths in China from Yu et al., in the following age classes: 0-24, 25-34, 35-44, 45-54, 55-64, 65-74, 75+ (37).

#### 8. All emergency visits in France

We obtained the age and sex distribution of all coded emergency visits (all causes) in French hospitals from the OSCOUR® network between 2004 and 2019.

#### 9. All cause mortality in 28 European countries

We extracted the all-cause number of deaths by single year age class and by sex for the 28 EU countries (including the UK) through the Euromomo system.

#### 10. Multi-country SARS-CoV-2 mortality model estimates

We obtained estimates of the relative risk of death given infection by age and sex from a previously published model that fitted age and sex-specific COVID-19 death data from 45 countries(1).

### Data analyses

#### 1. Comparisons by age

We compared the incidence between each group using a rolling window of size 5 years with that of 60 year olds. Taking each pathogen in turn, we compared the incidence within each age group by dividing the total number of deaths within an age group with the size of the population from the national census for that country. We then calculated the relative incidence within each age group as compared to 60 year olds. This approach allows us to adjust for underlying differences in incidence and outcome across pathogens that affect all age groups in the same way. We considered three different outcomes: Hospitalisation (or emergency department visit in the case of influenza), severe disease (defined as requiring ICU treatment or ending in death) and death. For all cause emergency department visits, we removed the cases that were due to influenza to ensure any patterns weren’t being driven by influenza admissions. To capture uncertainty we calculated 95% Wald confidence intervals. For the multi-country SARS-CoV-2 mortality estimates, the risk of death given infection was derived from a Bayesian model that combined the results of 22 national-level seroprevalence studies with reported age and sex-specific COVID-19 death data from 45 countries. Relative risks by age were calculated in 5 year age groups as compared to those aged 60-64 years.

#### 2. Comparisons between males and females

For all-cause hospitalisation and mortality, influenza hospitalisation (France) and death (USA), and SARS-CoV-2 hospitalisation (France), severe disease (France) and death (multicountry study), sufficient data was available to detect differences by both sex and age. We compared the incidence between males and females using a rolling window of size 10 year age groups (5 year groups in the case of the multi-country SARS-CoV-2 analysis). Taking each pathogen in turn, for each window, we divided the incidence in males with that observed in females. We calculated 95% Wald confidence intervals.

---

**Figure S1.**
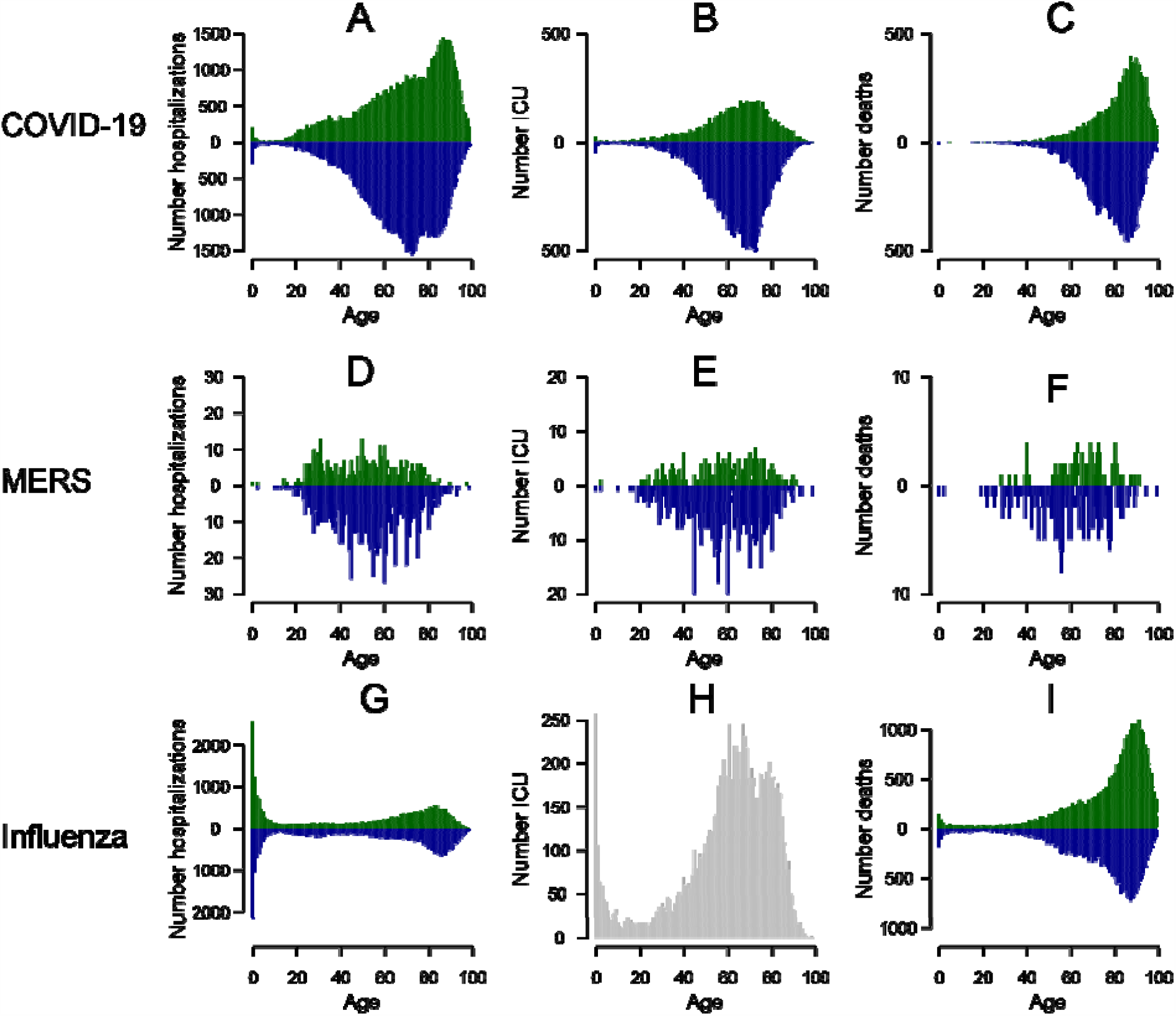
Case distributions for COVID-19, MERS and Influenza. **(A)** COVID-19 hospitalizations in France by age comparing females (green) and males (blue). **(B)** COVID-19 hospitalizations requiring ICU in France by age comparing females (green) and males (blue). **(C)** COVID-19 deaths in France by age comparing females (green) and males (blue). **(D)** Total MERS hospitalizations by age comparing females (green) and males (blue). **(E)** Total MERS hospitalizations requiring ICU by age comparing females (green) and males (blue). **(F)** Total MERS deaths by age comparing females (green) and males (blue). **(G)** Total influenza hospital emergency visits in France (2004-2020) by age comparing females (green) and males (blue). **(H)** Total hospital visits for influenza requiring ICU in France (2012-2017) by age. **(I)** Total hospital influenza deaths in US (2019) by age and sex.

**Figure S2.**
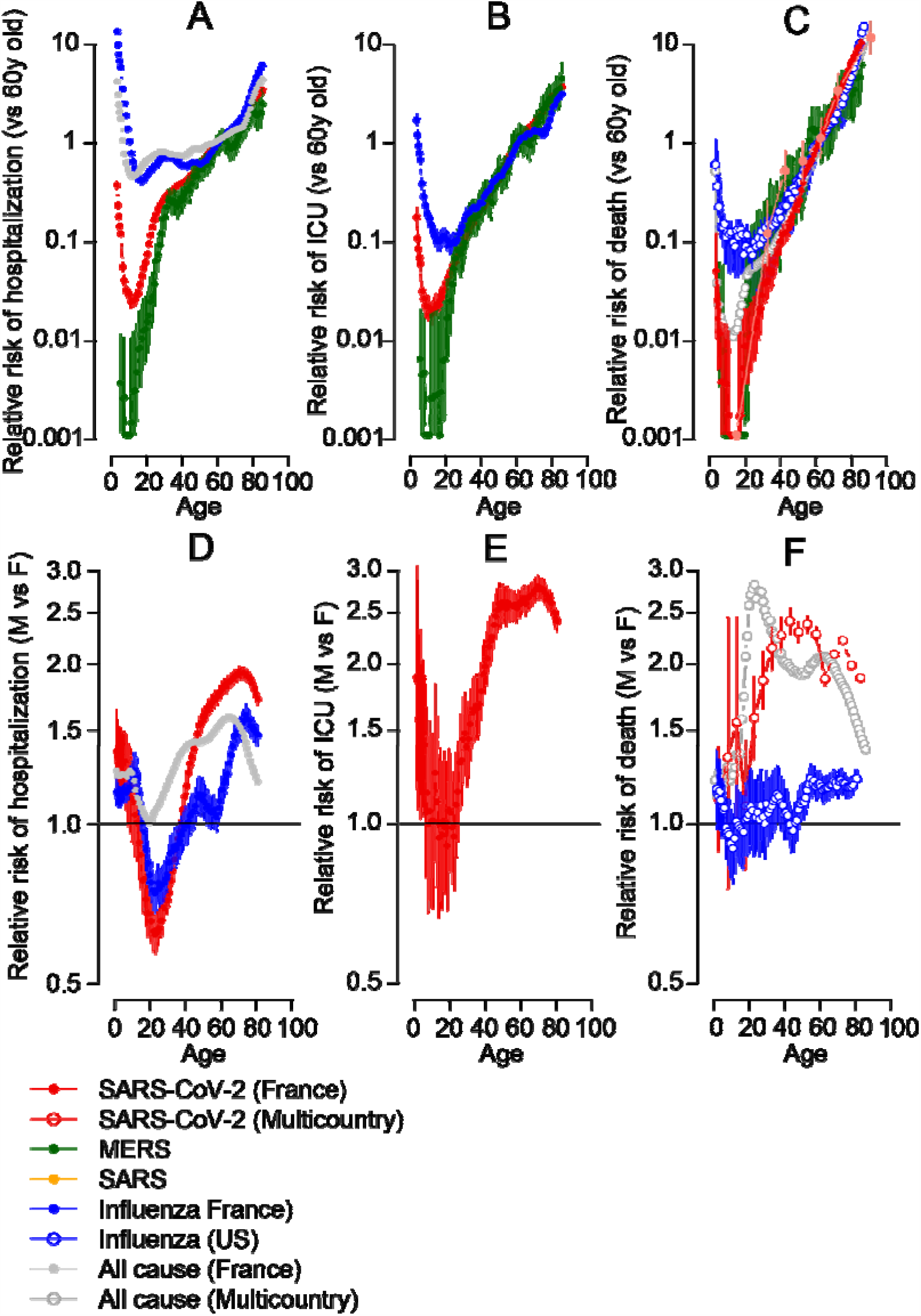
Relative risk of hospitalisation, severe disease and death, including 95% confidence intervals.

**Figure S3.**
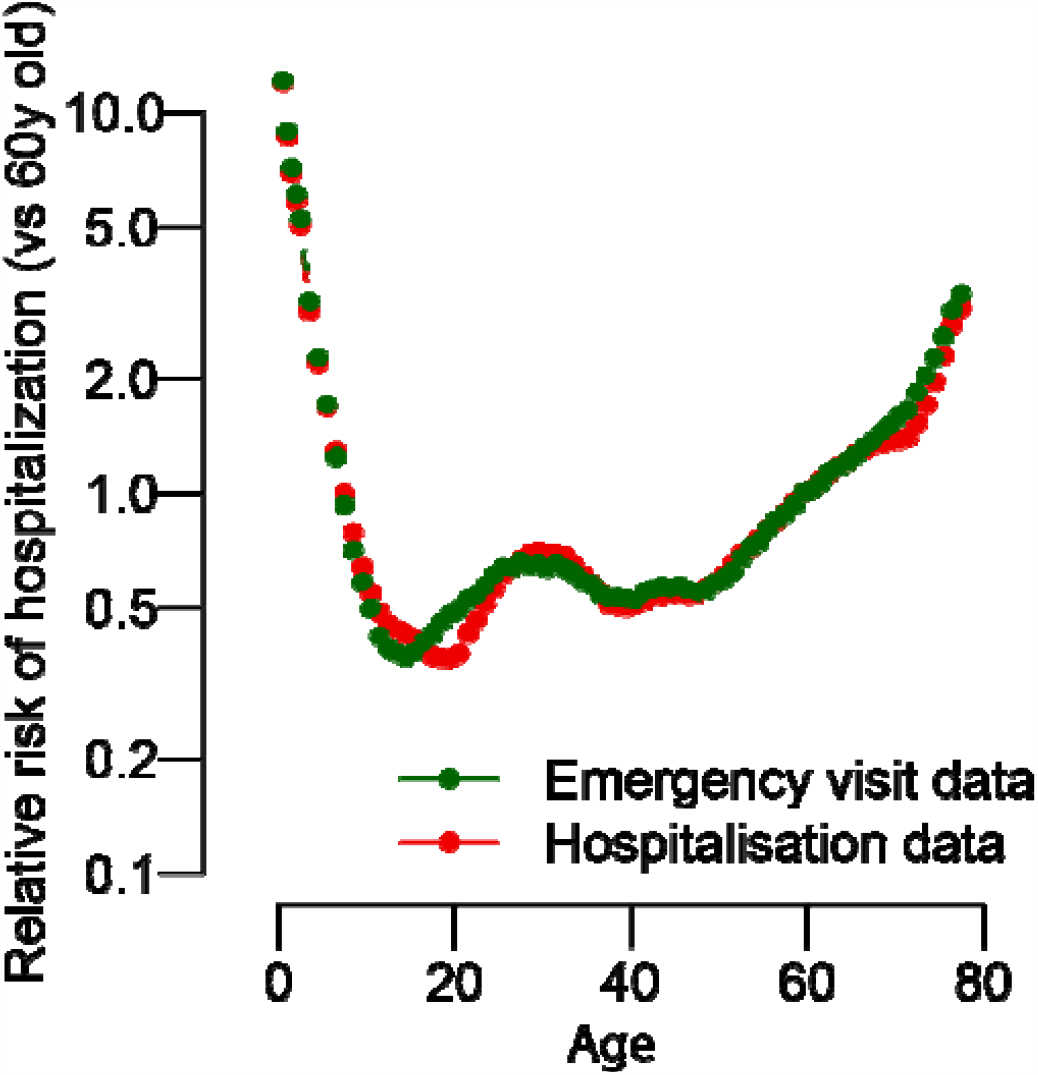
Relative risk of hospitalization by age as compared to a 60y old for influenza in France. Comparison between emergency visits for influenza (green) between 2004 and 2019 and all hospitalisations for influenza (red) between 2012 and 2019.

**Figure S4.**
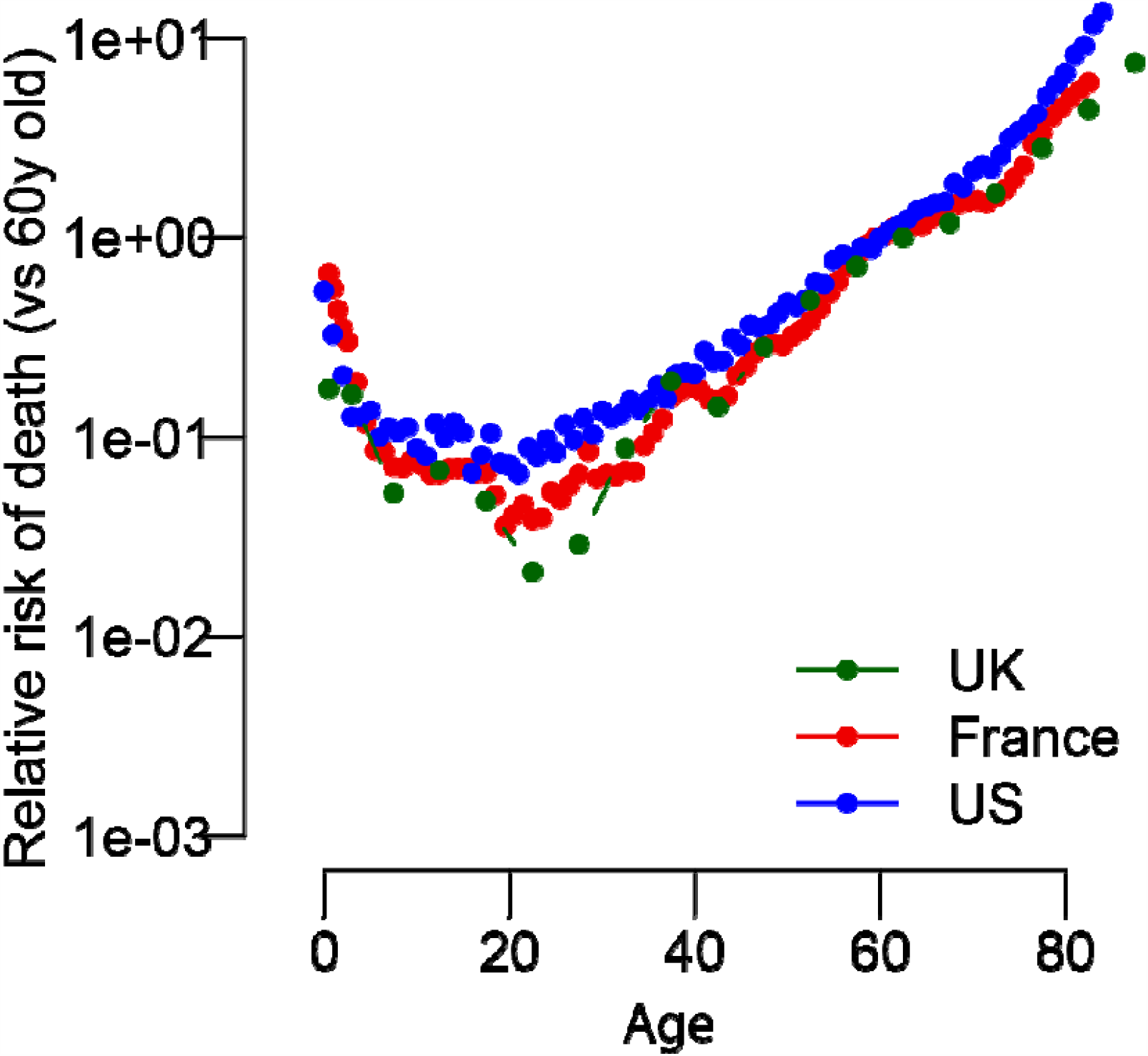
Relative risk of death from influenza for France, US and UK.

